# CLINICAL APPLICATIONS OF MACHINE LEARNING ON COVID-19: THE USE OF A DECISION TREE ALGORITHM FOR THE ASSESSMENT OF PERCEIVED STRESS IN MEXICAN HEALTHCARE PROFESSIONALS

**DOI:** 10.1101/2020.11.18.20233288

**Authors:** Juan Luis Delgado-Gallegos, Gener Avilés-Rodriguez, Gerardo R. Padilla-Rivas, María De los Ángeles Cosio-León, Héctor Franco-Villareal, Erika Zuñiga-Violante, Gerardo Salvador Romo-Cardenas, Jose Francisco Islas

## Abstract

Stress and anxiety have shown to be indirect effects of the COVID-19 pandemic, therefore managing stress becomes essential. One of the most affected populations by the pandemic are healthcare professionals. Thus, it is paramount to understand and categorize their perceived levels of stress, as it can be a detonating factor leading to mental illness. In our study, we used a machine learning prediction model to help measure perceived stress; a C5.0 decision tree algorithm was used to analyze and classify datasets obtained from healthcare professionals of the northeast region of Mexico. Our analysis showed that 6 out of 102 instances were incorrectly classified. Missing two cases for mild, three for moderate and 1 for severe (accuracy of 94.1%), statistical correlation analysis was performed to ensure integrity of the method, in addition we concluded that severe stress cases can be related mostly to high levels of Xenophobia and Compulsive stress.

## Introduction

As the deadly coronavirus disease COVID-19 continues to spread globally medical and allied healthcare professionals have become one of the most highly affected sectors by this disease^1–3^. Particularly in developing democracies, such is the case of Mexico, the public health system has become engulfed by the overwhelming levels of stress^4,5^ In addition, the situation becomes even more taxing for attending personnel as they not only deal with the burdened system^6^, but they must also deal with the enemy upfront; It is here, where they too can become prey to the disease^7^. Recent reports showed for the period spanning from the end of February to the end of August 97,632 health-care professionals became infected with over 1300 more deaths reported over any other country^8^. What is even more burdensome is the fact that Mexico continues atop of all Latin-America countries in infection-to-death rate (>10%)^9^.

According to the Pan-American Health Organization (PAHO), Mexico has the highest number of healthcare workers with COVID-19 in Latin America^10^. Up to October 25^th^ 2020, the amount of healthcare professionals affected by COVID-19, as reported by the National health ministry, by comparison to other healthcare professionals in Mexico, medical doctors rank highest in mortality vs their nursing or other allied healthcare professional counterparts. Recent reports show that both physicians and nurses have similar levels of burnout and emotional fatigue. Physicians tend to work in a more independent manner, this along with their long shift hours, high-sense of duty, work ethics, and the fact they partake multiple jobs, normally of low wages, becomes a source of additional stress^8^. Counter, nursing staff work in groups and develop higher social support^11,12^. Recent reports have shown that resilience level in nurses seem to correlate with lower anxiety, this is part of a well-developed social support system in which nursing professionals provide emotional help and support in order to uplift the communal spirit even under overwhelming circumstances^13^.

During the period of August 16th up to November 3rd, the spreading of the disease amongst physicians grew significantly along the passing months, reporting 140,196 confirmed cases, 31,870 classified as “possible” COVID-19 and 222,372 discarded cases, with 1,884 COVID-19 confirmed deaths and up to 198 COVID-19 suspected deaths, with an active case count of 3362. Interestingly, active cases decreased from the initial count of 4743 in the period of August, to 3362, nonetheless in November the number of suspected and confirmed COVID-19 death rate showed an increase^14^ (Figure 1). This is of particular interest when we take into account stress, as it may be a trigger to determine a potential severe outcome of the disease.

**Figure 1.**
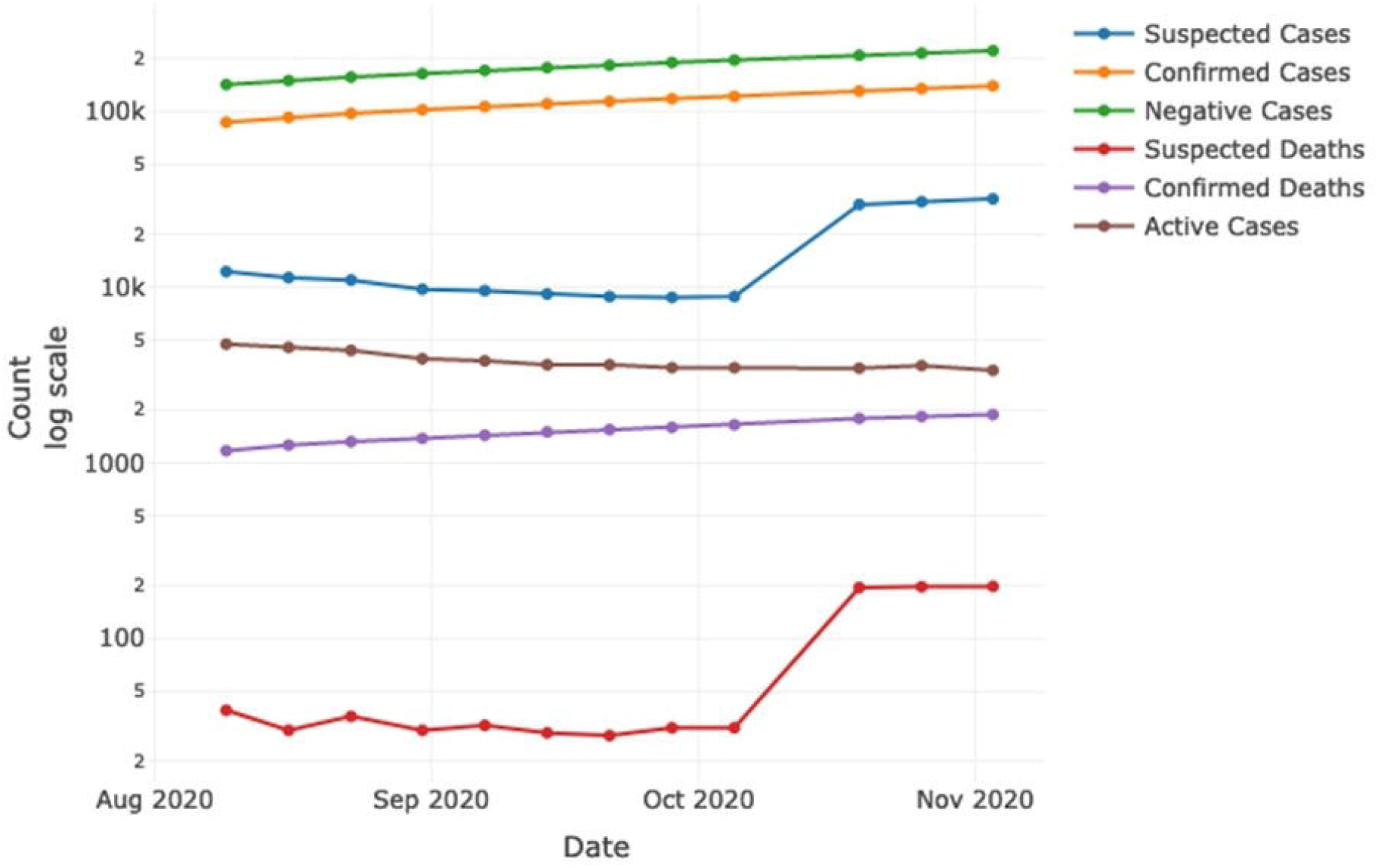
COVID-19 epidemiological curve for physicians in accordance to the Mexican health ministry form the periods of August to November 2020. Increases, confirmed cases (140,196), suspected cases (31,870), negative cases (222,372), confirmed deaths (1,884), suspected deaths (198). Decreases, active cases (3362).

Recent developments in computational modeling have led to the ever-evolving field of artificial intelligence which, when combined with neuro- and behavioral-science, has created the new field of computational psychiatry^15,16^. Nowadays Machine Learning and Artificial Intelligence are promising technology used by various healthcare providers as they result in better scale-up, speed-up processing power, reliability which translates into the strengthening of the performance of the clinical team^17^. Therefore, there’s has been an expected trend in using these techniques in order to better understand and fight against the pandemic. Computational psychiatry helps to model and understand underlying mental illness, when this is driven forward with “machine learning” we can potentially predict behavioral patterns, improve classification and assist the physician to provide a faster and personalized medical attention.

Well-known machine learning algorithms are decision trees, commonly used for establishing classification systems based on multiple variables for a target feature^18^. Typically, with the use of this method, it is possible to classify a population into branch-like segments that generate an inverted tree^18,19^. The algorithm can efficiently deal with large, complicated datasets without imposing a complicated parametric structure^19^. Researchers have reported the use of these types of algorithms for applications in the study of behavioral and mental health^20^. Thus, it is possible to use this technique to help define different clinical paths, classify subgroups of patients for different diagnostic tests, treatment strategies, and assessment of mental health-related conditions^21,22^.

## Methods

A common strategy used for data analytics is the Cross Industry Standard Process for Data Mining methodology (CRISP-DM), for which six steps are defined for data-based knowledge projects. This strategy begins with a phase where problems from the scenario are stated and objectives defined (Business Understanding), followed by a stage where data insights are obtained (Data Understanding). Next, a final dataset for study is defined and analyzed (Data Preparation), results from the data preparation analysis would allow to define the algorithm over which a model would be generated (Modeling) once the model is generated a performance evaluation would be required in order to confirm if it fulfills the proposed objectives (Evaluation), if the goal for which the modeling is achieved, it can be implemented for the purpose for which it was proposed^29^.

This work is based on previously reported adapted COVID-19 stress scales data^1^. The analysis method was adapted from CRISP-DM, in the same amount of stages and sequence, as shown in Figure 4. An initial Data Structure study was done from the data analytics scope in order to consider the type of variables that compose the stress scales. Also, to define the goal for the study as a model that contributes in the understanding of mental illness in health care workers during the COVID-19 outbreak. Next, a Data Validation analysis that considered statistical test to confirm relations between variables from the scales and classification outcomes from raw data, and also, to confirm internal consistency. This was followed by a Data Distribution analysis to study stress components that could be used for model selection and interpretation. A Decision tree model was selected in order to study the relations and possible classifications routes for stress level according data from its respective scales. Given the clinical component of the study, a performance analysis based on accuracy, sensitivity and specificity was carried out on the results. This allowed to validate the obtained model and Observations and conclusions from stress scales were made as an application of the obtained model.

**Figure 2.**
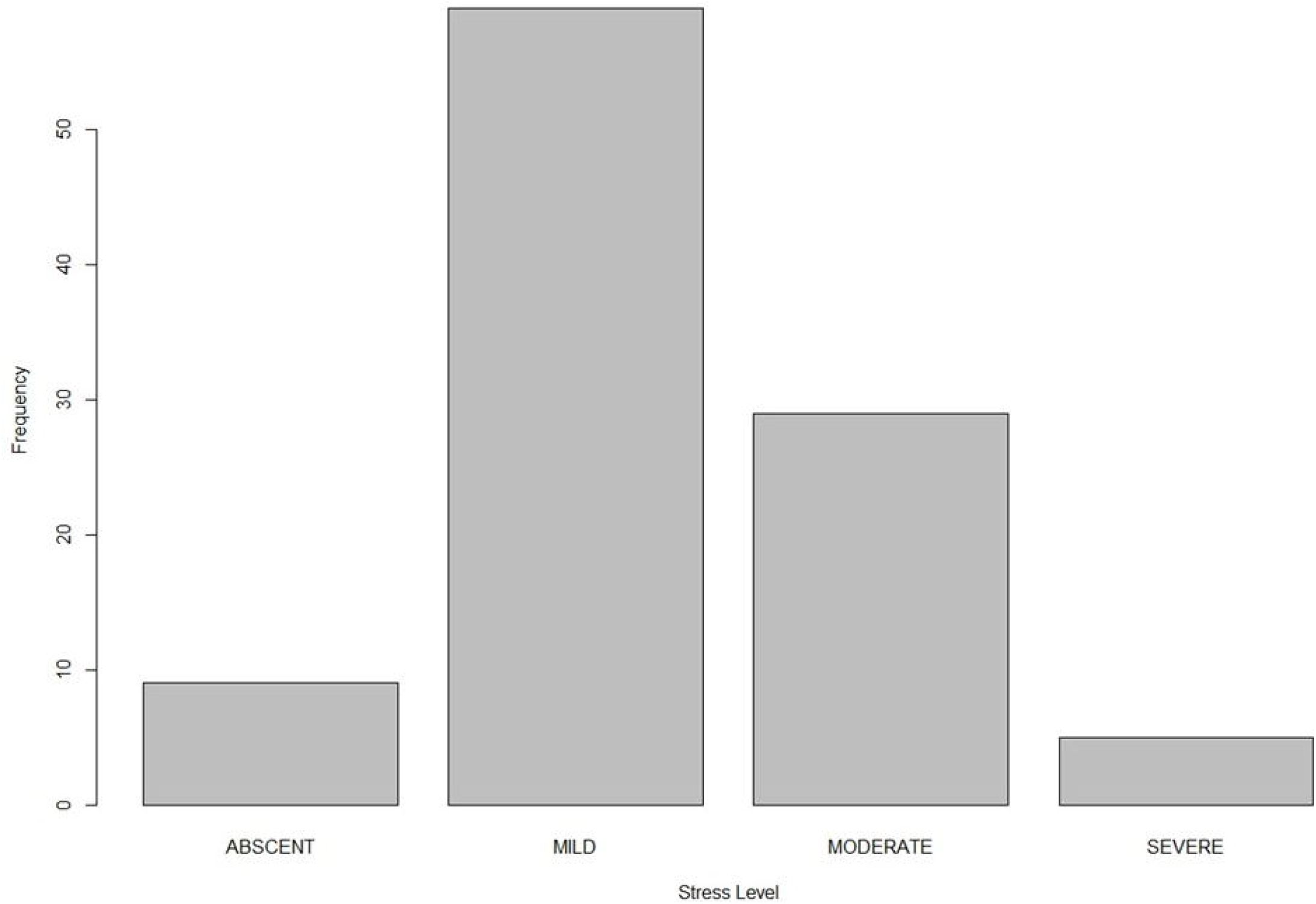
Stress level distribution in Healthcare personnel. (Left to right) Absent, Mild, Moderate, Severe.

**Figure 3.**
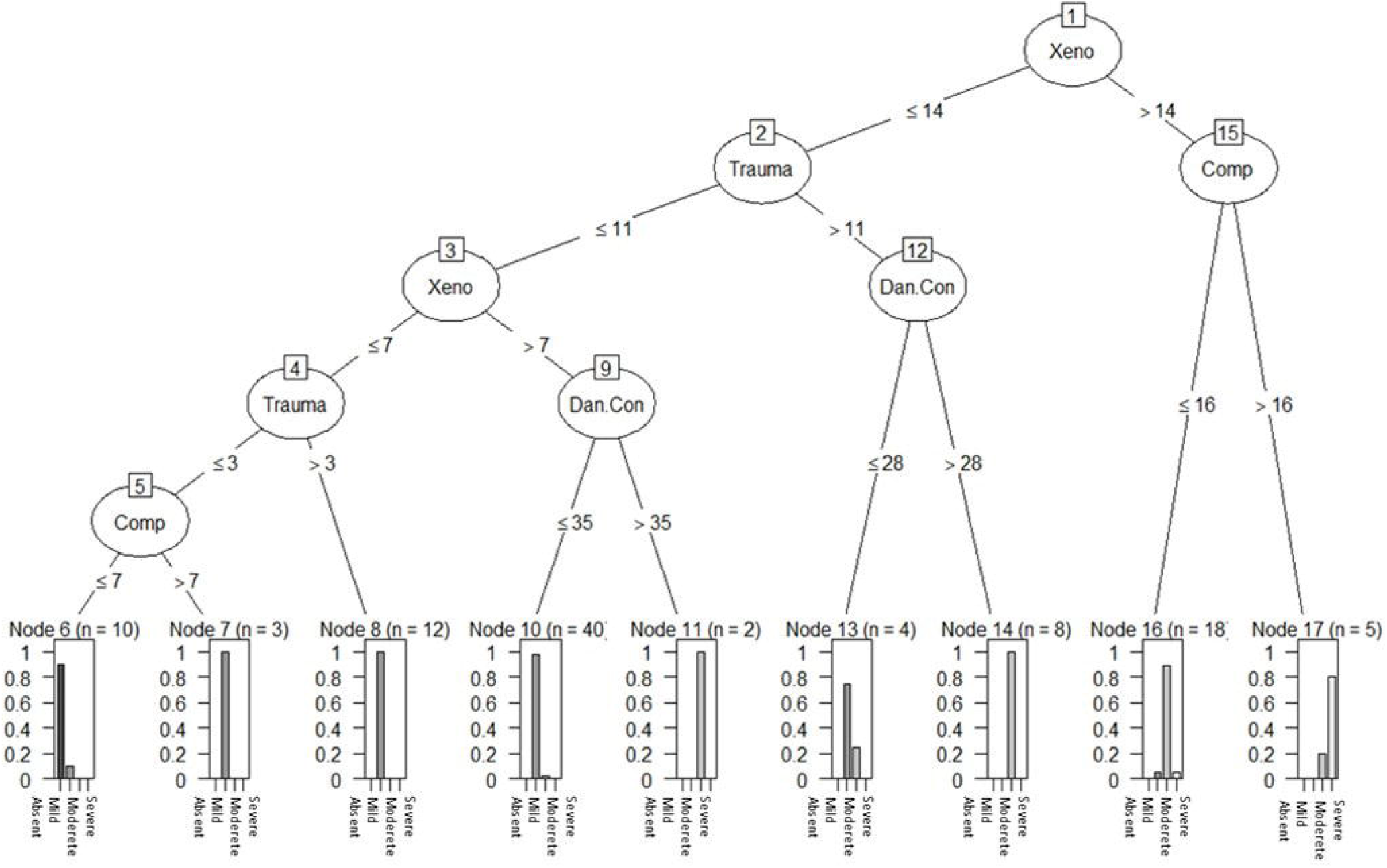
Decision tree applied into healthcare personnel stress scale level dataset. Atop variables influencing stress are Xenophobia (Xeno) and Compulsive checking (Comp), which leads to severe stress. Traumatic stress (Trauma) and Danger + Contamination (Dan Con) also influenced the perception of stress. Social economical variable did not influence the outcome of the decision tree.

**Figure 4.**
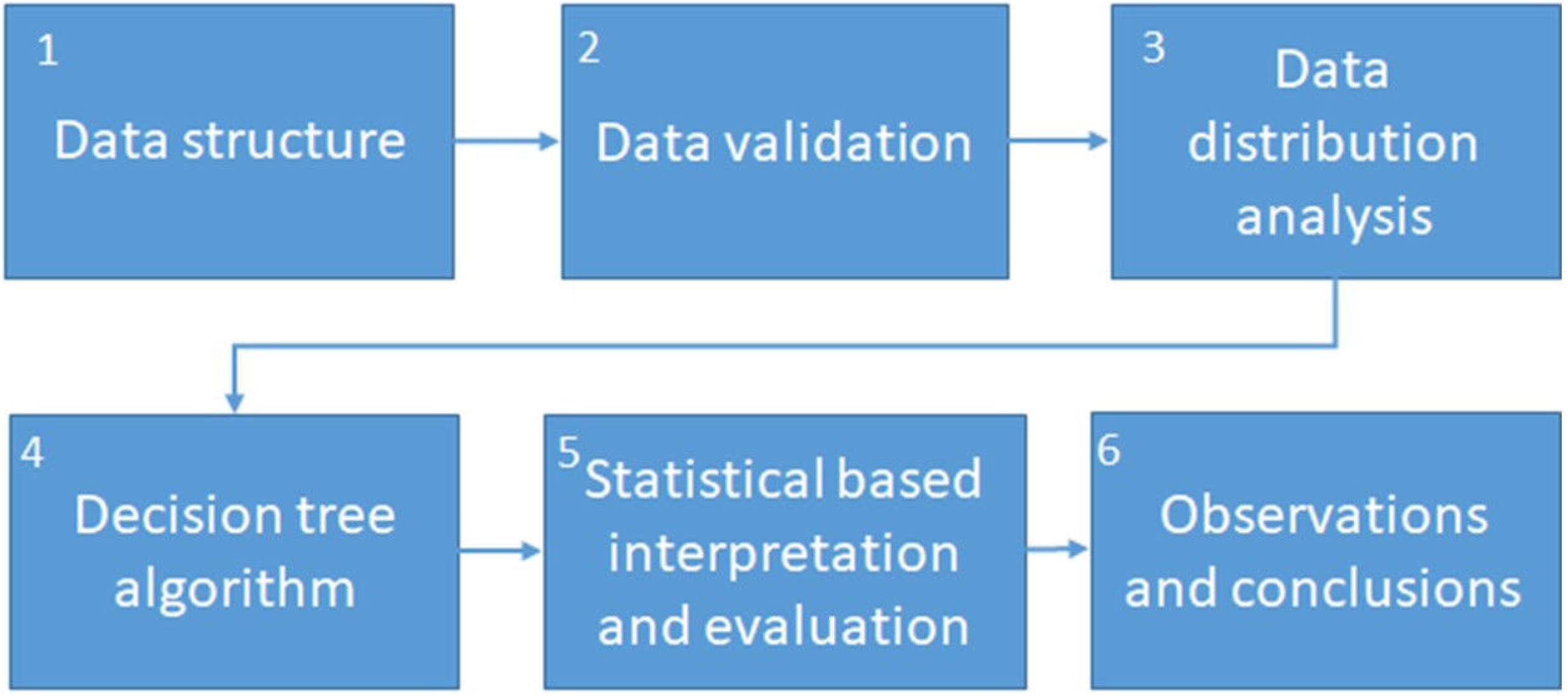
Methods for machine learning based analysis on stress scales of health care workers.

The statistical analysis and computational modeling (algorithm) were done to obtain the behavior patterns and distribution of the data. Both the statistical and algorithm analysis of the dataset was performed in R and RStudio.

### Descriptive statistical analysis

In order to understand the structure, validate and explore data distribution, a descriptive statistical study was carried out. The comprised obtaining measures of central tendency and variability metrics to describe the whole-set of data with a single values, the center of distribution and similitude^30^. Cronbach’s alpha was estimated considering the numerical values from all participant responses. Presented as the index of reliability and internal consistency^31^. Chi-square statistic was applied to the general classification of each participant and each of the questions separated by the sections of the generalized stress scale reported^32^.

### Mathematical and computational algorithms

Following the statistical analysis that implied the first three stages of the study. A decision tree model was developed to behave as a computational supportive scaffold for the study of mental illness. These types of algorithms have previously been useful to study and predict mental illness^19,20^. C5.0 algorithm was used to analyze and classify the stress level from the dataset^23^, and for construction of decision tree algorithm^23^. The relation between entropy and information gain allows for a model, that trains based on the datasets that contribute to analyze and visually explain the route and relations of the studied variables. Excluding features that don’t affect the outcome. Making it an efficient tool for the understanding of illness and its severity based on measures stress features.

Following both the statistical and computational study of the dataset, performance based on sensibility, sensitivity and accuracy was studied on the generated model^33^. Conclusions were drawn from the outcome and routes defined by the tree model branches considering initial statistical analysis.

## Results

An initial preprocessing statistical analysis was applied to the 106 instances dataset. After eliminating missing data instances for statistical and algorithm-based analysis preparation, a group of 102 instances was used for the study.

Frequency counts were applied to the Participant profession and work area variables, as shown in Table 1.

**Table 1.**
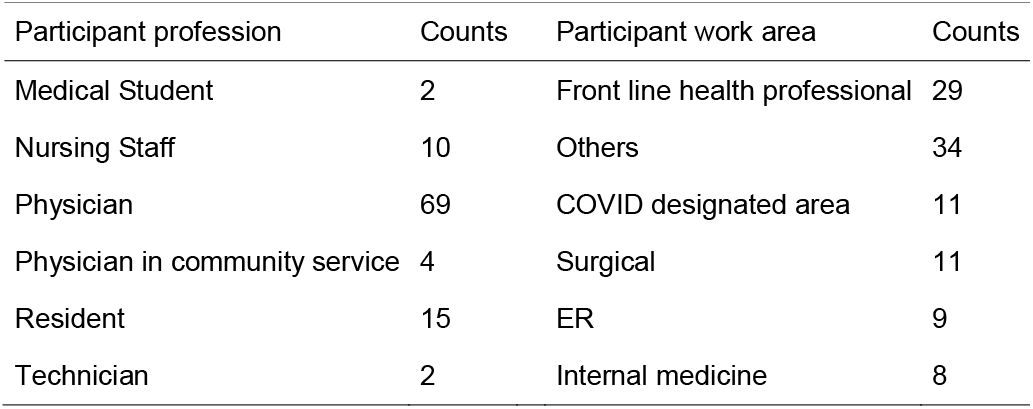
Frequency count on Participant profession and work area.

The adapted COVID-19 stress scales were built upon five areas that come because of the addition from the response of the questions related to each section of the survey. Central tendency metrics were calculated for each of these components based on the cumulative result of each participant, as shown in Table 2.

**Table 2.** Central tendency metrics for the adapted COVID-19 stress scale features..

| Stress scale feature | Min | 1st Quartile | Median | Mean | 3rd Quartile | Max |
| --- | --- | --- | --- | --- | --- | --- |
| Danger and Contamination fear | 5 | 23 | 25 | 25.2 | 33.75 | 48 |
| Socioeconomical | 4 | 14 | 17 | 16.27 | 19 | 24 |
| Xenophobia | 1 | 7 | 10.5 | 10.9 | 14 | 24 |
| Traumatic stress | 0 | 2 | 6 | 7.37 | 12 | 22 |
| Compulsive checking | 0 | 5 | 8 | 9.38 | 13.75 | 24 |

Given that each feature is based on the addition of the responses from the survey. We considered all the values from each question and participant for the calculation of Cronbach’s alpha. This gave a result of 0.94 which shows a good internal consistency for the whole survey instrument and data. In addition, supplementary Table 1 shows Chi-square tests to each question for all in order to define significance in the relationship of the variables. Table 3 shows the result of the test for each scale area and each question and for the cumulative adapted COVID-19 stress scales.

**Table 3.** Analysis per General Area.

| COVID areas | absent | mild | moderate | Severe | Xi2 | sig |
| --- | --- | --- | --- | --- | --- | --- |
| Danger + Fear of Contamination | 3 | 23 | 58 | 17 | 64.98 | <0.001 |
| Socioeconomical | 30 | 35 | 24 | 12 | 11.673 | <0.009 |
| Xenophobia | 15 | 45 | 29 | 12 | 27.119 | <0.001 |
| Traumatic stress | 47 | 25 | 21 | 8 | 31.238 | <0.001 |
| Compulsive checking | 26 | 43 | 22 | 10 | 22.129 | <0.001 |
| CSS general score | 9 | 59 | 28 | 5 | 72.109 | <0.001 |

Xi2= Chi-square test

Most of the results from the Chi-square test show significance, except for the following: one question from the Traumatic stress scale and four from Compulsive checking scale. This relating dependence of the stress level classification calculated as a cumulative result of the scales and the answers given by each participant.

The distribution for the stress level classification in healthcare personnel calculated from the stress scales is shown Figure 2. Frequency histogram of the four severity levels of the stress scale from the observed dataset. This variable would correspond to the target feature in the decision tree model (Figure 3).

Both results from the Cronbach alpha and the Chi-square test, show internal consistency of the data and validate the dependence for stress level calculation, ensuring the dataset quality for algorithm-based analysis.

Following the descriptive statistical analysis, a decision tree model was trained with the preprocessed dataset using the C5.0 algorithm^19,23^. Considering the stress level to be the target variable. Participant profession, work area and all five cumulative stress scales areas were used as the predictive variables in order to find any relationship between them aiming to predict stress level. Figure 3 shows the decision tree obtained from the dataset. At the predictive branch, a set of boxes with all four levels of stress are observed. In each box, the extreme right bar corresponds to the severe level indicator, followed to the left by moderate, mild and absent levels, respectively, also observed in Figure 3. Despite declaring the features related to participant profession and work area, these variables were pruned from the model. Table 4 shows the confusion matrix from the obtained decision tree model. Where only 6 out of 102 instances were incorrectly classified. Missing two cases for mild level, three for moderate and 1 for severe. All these errors were classified only in neighboring levels, giving the model an accuracy of 94.1%.

**Table 4.** Confusion matrix of obtained decision tree model for stress level classification.

| Classified as |  |  |  |  |
| --- | --- | --- | --- | --- |
| a) | b) | c) | d) |  |
| 9 |  |  |  | a) |
| 1 | 57 | 1 |  | b) |
|  | 2 | 26 | 1 | c) |
|  |  | 1 | 4 | d) |

a) absent b) mild, c) moderate, d) severe

In order to analyze model performance, a sensibility and specificity calculation was carried out. For this, three different scenarios were considered based on the classification outcome from the dataset, dividing instances in healthy and disease groups. Calculation was done with the figures from the confusion matrix. Results shown in Table 5.

**Table 5.**
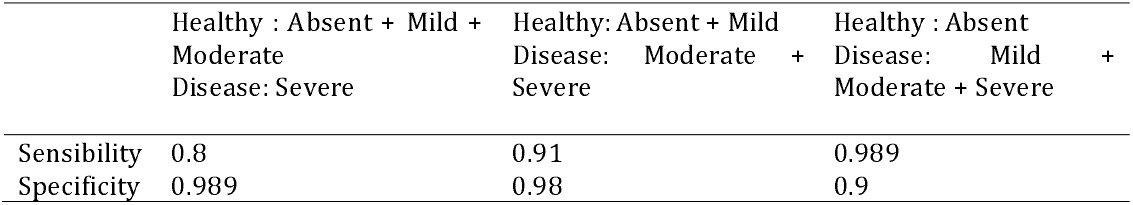
Decision tree sensitivity and specificity calculation from stress scales dataset.

## Discussion

Our purpose was to define a statistical and computational framework algorithm to understand stress levels in healthcare professionals due to the impact of the COVID-19 pandemic and to potentially define a tool which can be further used as a predictor of severity of stress.

A dataset related to adapted COVID-19 stress scales as defined by Delgado-Gallegos *et*.*al*., was studied with a calculated Cronbach alpha of 0.94 which shows a good internal consistency; stress levels were calculated as a geometrical result from the addition of five scales from the survey defined as Danger + Fear of contamination, Socioeconomic stress, Xenophobia, Traumatic stress and Compulsive checking. Chi-square test was done for all questions individually, looking to validate stress level calculation. Statistical significance (p < 0.05) was found in most of the questions considering the answer of all participants, except one question in the Traumatic scale, and four from the Compulsive checking scale (all shown in supplementary table1), although all scales showed statistical significance when the test was applied to the accumulated value for each of these scales, as seen on Table 3. Thus validating, the use of the adapted COVID-19 stress scales, in a population^1,24^. Recently the Mexico’s health ministry has also launched their own questionnaires, in an effort, to correctly assess the stress levels in healthcare professionals^25^. Therefore, the use of this model can be re-adapted to help in correctly assessing and provide a faster diagnosis and opportune treatment.

From the central metrics statistical analysis, no relation was observed between participant profession and work area, similar analysis was done for the stress scales which showed an exception for Danger + Fear of contamination joint scale, all other areas had a similar maximum value but with different means. Therefore, considering results from the preprocessing stage, the dataset shows good quality, independence and internal consistency for algorithm analysis. All 102 instances from the dataset were used to train a decision tree model by C5.0 algorithm, where stress level was defined as the target variable, with participant profession, work area and cumulative stress scales as predictors. The resulting model showed an accuracy of 94%. Nonetheless, the algorithm did not find enough information gain from the participant profession, work area, and the socioeconomic scale. Neglecting these variables from the resulting model allows to understand that experience and day-to-day work routine are not a factor on how healthcare professionals perceive stress. Resilience could help explain this pattern as it is an adaptation mechanism in which a person overtime can handle stress in overwhelming situations ^13,26^.

## Conclusion

Computational psychiatry states the similarity between the brain and a computer and proposes the use of computational terminology for the study of mental illness^27^. Our results show interesting data denoting hypothetical tendencies based on the purity of the resulting branches of the decision tree, where severe stress cases can be related mostly to high levels of Xenophobia and Compulsive stress. This considering that threshold values from the extreme right route of the decision tree are above the 3rd quartile for both scales. In a similar manner, absent stress level comes from the scenario of combined thresholds below the 1st quartile from Xenophobic, Compulsive and Traumatic stress scales. It is interesting to note that the Danger + Fear of contamination scale can be used to find both mild and moderate cases, despite being a larger joint scale.

We believe this method could potentially define concepts on how to diagnose and handle severe stress, contributing to mathematical informed understanding of mental illness and computational psychiatry, thus forming a diagnostic tool to help in the assessment of patients. In this study, we used healthcare professionals as they are one of the most affected sectors in the pandemic^28^. In addition, an expansion of this method outside the pandemic can be used to understand different stress factors and how they can interfere with performance and social dynamics in different populations.

While this is only a fist approximation based on recent data from healthcare professionals in the northeast part of Mexico^1^, the impact of applying machine learning algorithms and computational psychiatry, can potentially help reshape the way we run global health.

## Data Availability

DATA AVAILABILITY: Dataset may be downloaded from Kaggle (https://www.kaggle.com/chepox/css-mexico).
CODE AVAILABILITY: Code may be accessed from github (https://github.com/Bio-Math/COVID-19-related-stress-analysis)

https://github.com/Bio-Math/COVID-19-related-stress-analysis

## DATA AVAILABILITY

Dataset may be downloaded from Kaggle (https://www.kaggle.com/chepox/css-mexico).

## CODE AVAILABILITY

Code may be accessed from github (https://github.com/Bio-Math/COVID-19-related-stress-analysis)

## Author contributions

Research and writing, J.L.D.-G., J.F.I., G.A-R, E.Z-V and H.F.-V.; algorithm development, M.D.A.C-L., G.S.R-C, G.A-R., statistical analysis, J.L.D.-G. and G.R.P.-R.; supervision, J.F.I. All authors have read and agreed to the published version of the manuscript.

## Figures and supplemental legend

**Supplementary Table 1**. Proportional analysis per question and overall area, Chi-square Test, Sig.

