## supplemental Table 1 for "CLINICAL APPLICATIONS OF MACHINE LEARNING ON COVID-19: THE USE OF A DECISION TREE ALGORITHM FOR THE ASSESSMENT OF PERCEIVED STRESS IN MEXICAN HEALTHCARE PROFESSIONALS"

SUPPLEMENTAL DATA

Supplementary Table 1. Proportional analysis per question and overall area, Chi-square Test, Sig.

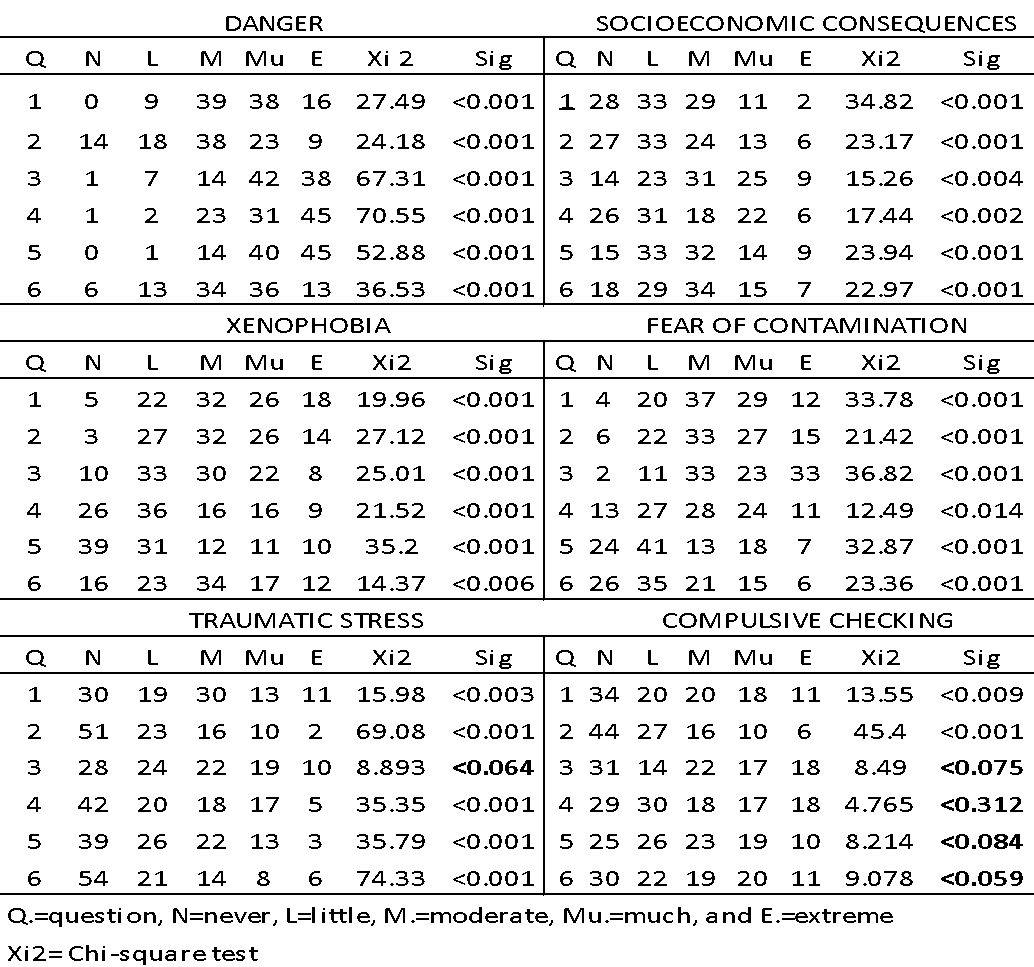
